# Cohort Profile: The United Kingdom Research study into Ethnicity And COVID-19 outcomes in Healthcare workers (UK-REACH)

**DOI:** 10.1101/2022.01.19.22268871

**Authors:** Luke Bryant, Robert C Free, Katherine Woolf, Carl Melbourne, Anna L Guyatt, Catherine John, Amit Gupta, Laura J Gray, Laura Nellums, Christopher A Martin, I Chris McManus, Claire Garwood, Vishant Modhawdia, Sue Carr, Louise V Wain, Martin D Tobin, Kamlesh Khunti, Ibrahim Akubakar, Manish Pareek, the UK-REACH Collaborative Group

**Author notes:** Joint 1^st^ Authors (Luke Bryant, Robert C Free). Manish Pareek (Chief investigator), Laura Gray (University of Leicester), Laura Nellums (University of Nottingham), Anna L Guyatt (University of Leicester), Catherine John (University of Leicester), I Chris McManus (University College London), Katherine Woolf (University College London), Ibrahim Akubakar (University College London), Amit Gupta (Oxford University Hospitals), Keith R Abrams (University of York), Martin D Tobin (University of Leicester), Louise Wain (University of Leicester), Sue Carr (University Hospital Leicester), Edward Dove (University of Edinburgh), Kamlesh Khunti (University of Leicester), David Ford (University of Swansea), Robert Free (University of Leicester).

## Abstract

Key Features of the UK-REACH Cohort (Profile in a nutshell)

- The UK-REACH Cohort was established to understand why ethnic minority healthcare workers (HCWs) are at risk of poorer outcomes from COVID-19 when compared to their white ethnic counterparts in the United Kingdom (UK). Through study design, it contains a uniquely high percentage of participants from ethnic minority backgrounds about whom a wide range of qualitative and quantitative data has been collected.
- A total of 17891 HCWs aged 16-89 years (mean age: 44) have been recruited from across the UK via all major healthcare regulators, individual National Health Service (NHS) hospital trusts and UK HCW membership bodies who advertised the study to their registrants/staff to encourage participation in the study.
- Data available include linked healthcare records for 25 years from the date of consent and consent to obtain genomic sequencing data collected via saliva. Online questionnaires include information on demographics, COVID-19 exposures at work and home, redeployment in the workforce due to COVID-19, mental health measures, workforce attrition, and opinions on COVID-19 vaccines, with baseline (n=15 119), 6 (n=5632) and 12-month follow-up data captured.
- Request data access and collaborations by following documentation found at https://www.uk-reach.org/main/data_sharing.

## Why was the cohort set up?

UK-REACH is a UK-wide prospective cohort established in November 2020 in response to the COVID-19 pandemic (1). COVID-19 has spread rapidly across the world, causing significant levels of morbidity and mortality, and devastated health economies in many countries. Healthcare workers (HCWs) have been at the forefront of the response to the pandemic and thus have been identified as being at increased risk of infection by SARS-CoV-2 and associated adverse outcomes (2-4). Furthermore, a number of studies have indicated that this risk of infection and adverse outcomes is greater for individuals from ethnic minority groups, particularly when compared to white HCWs (3). Emerging evidence also suggests that ethnic minority groups may be at an increased risk of long-term COVID-19 sequelae and poor mental health outcomes such as anxiety, depression and post-traumatic stress. (5, 6).

The quality of data related to COVID-19 risk and outcomes in HCWs is relatively poor, with very few large-scale representative studies in clinical or ancillary HCWs in healthcare settings stratified by ethnicity or occupation type, once potential confounders have been controlled for. The UK-REACH longitudinal cohort aims to address this disparity by examining differences in COVID-19 clinical outcomes (diagnosis, hospitalisation, ICU admission), professional roles and well-being among ethnic minority and white HCWs. The cohort will study the impact on COVID-19 on physical and mental health of ethnic minority HCWs compared to white HCWs in the short and long term with consent for linkage with electronic health records for up to 25 years from the date of consent.

## Who is in the cohort?

Recruitment to the cohort began on 4^th^ December 2020, and continued until 28^th^ February 2021. In total, **17891** HCWs from across the United Kingdom have been recruited into the study. Participants were considered eligible for the study if they were over the age of 16 years, lived and worked in the UK, and worked in health and social care or were a member of one of the UK healthcare regulators. This included ancillary workers such as cleaners and porters in healthcare settings. HCWs were invited to participate through two different channels. One was via an invitation from the various healthcare regulators and membership bodies within the UK; while the other was through a selection of NHS trusts and health boards throughout the UK. A total of 12280 participants were recruited through the first route, with 1018 participants recruited through the second route, and the remaining 4593 recruited by visiting the study website directly or via social media.

A total of 1,052,875 email invitations were sent by the healthcare regulators and membership bodies, summarised in table 2. Twenty-nine NHS bodies consisting of NHS trusts in England, NHS regions in Scotland and NHS health boards in Wales (summarised in table 1) engaged with their staff to increase recruitment, with invitations placed in trust-wide emails detailing recent events and news in each respective trust.

**Table 1:**
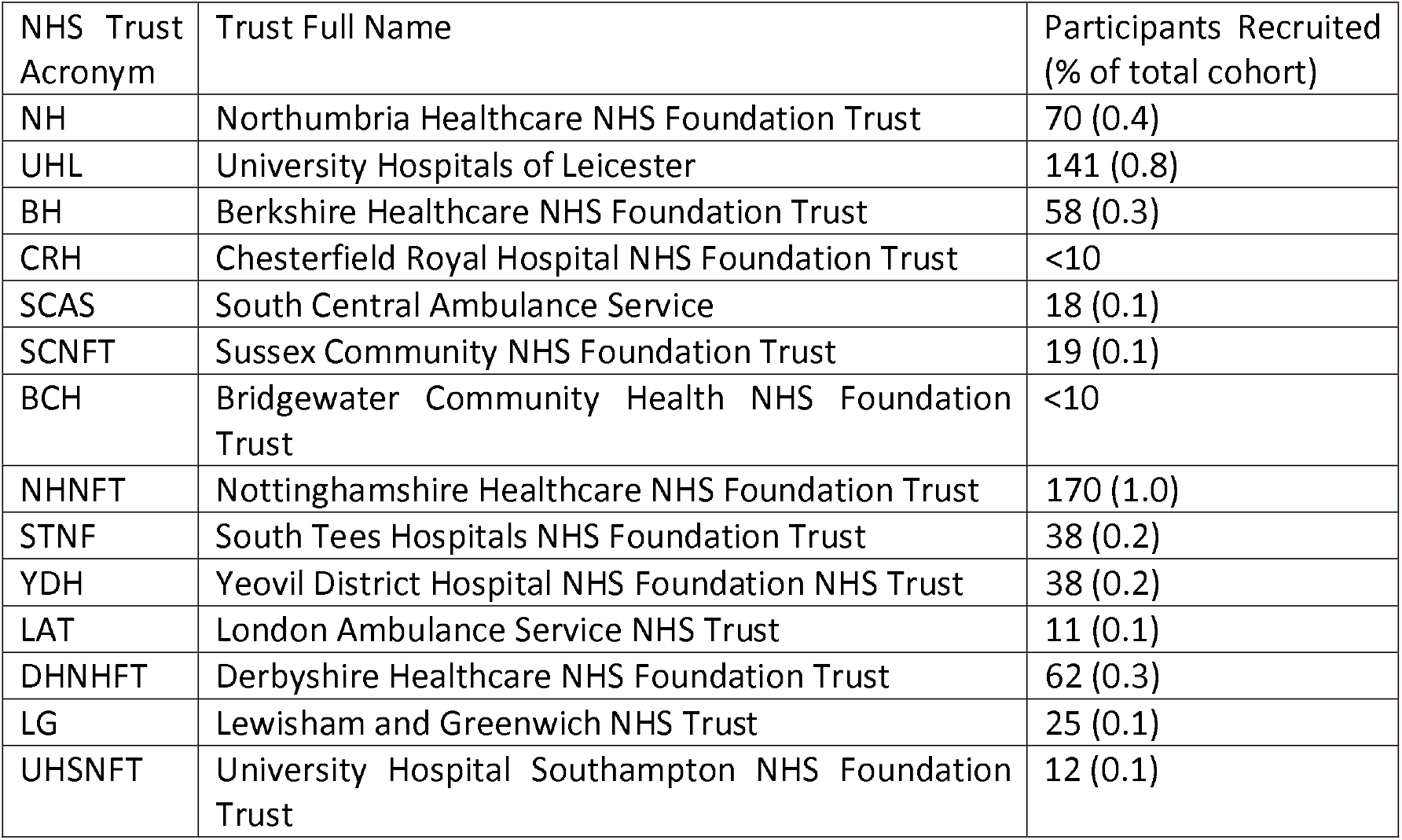
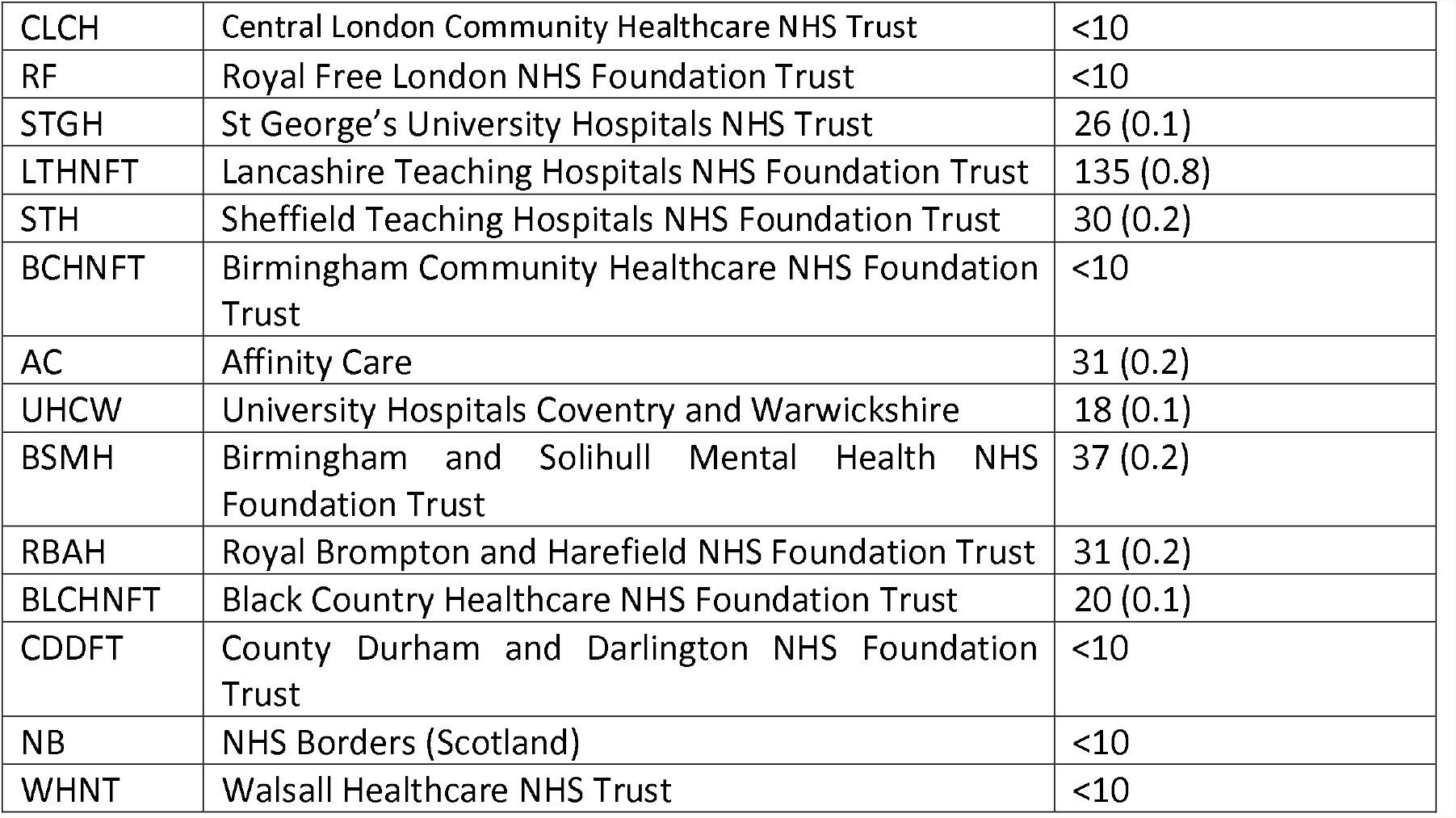
Recruitment from NHS Trusts and Health Boards into UK-REACH cohort. Trusts and Health Boards that recruited fewer than 10 participants have had their numbers masked to reduce the risk of participant identification.

**Table 2:** Participating Healthcare Regulators and Organisations in UK-REACH Cohort. Organisations who recruited fewer than 10 participants have had their numbers masked to reduce the risk of participant identification.

| Partner Acronym | Partner Full Name | Participants Recruited (% of UK-REACH cohort) |
| --- | --- | --- |
| GPhC | General Pharmaceutical Council | 212 (1.2) |
| GMC | General Medical Council | 3431 (19.2) |
| PSNI | Pharmaceutical Council of Northern Ireland | 27 (0.2) |
| GOC | General Optical Council | 344 (1.9) |
| NMC | Nursing and Midwifery Council | 2391 (13.4) |
| HCPC | Health and Care Professionals Council | 4963 (27.7) |
| GDC | General Dental Council | 905 (5.1) |
| Serco | Serco | <10 (0.03) |
| Unknown | Not through any recruiting site | 4593 (25.7) |

Interested participants were directed to the cohort website (https://www.uk-reach.org) where they were able to provide contact details along with informed electronic consent, including permission to link to Electronic Healthcare Records (EHRs) and to share pseudonymised research data with external researchers and to consent to participation in prize draws. The prize draw was offered to participants to incentivize completing individual questionnaires. Each prize draw consisted of 10 £250 amazon gift vouchers, 10 £50 amazon gift vouchers and 250 £5 amazon gift vouchers, taken after each questionnaire period was closed. In order to be eligible for each prize draw, participants were required to complete the most recent questionnaire.

## What has been measured?

After consenting to join the cohort, participants were invited to complete the baseline questionnaire, which addresses a range of topics related to COVID-19, leading to a wide range of qualitative and quantitative data being collected. A summary of the topics covered in the baseline questionnaire can be found in table 3.

**Table 3:** Summary of data collected at each phase.

| Phase (Dates) (n) | Topics (1) |
| --- | --- |
| Baseline Questionnaire (December 2020-February 2021) (n=15471) | Ethnicity<br>Nationality, religion and languages<br>Other demographics and education<br>Work<br>Home and Social Life<br>Harassment and discrimination<br>Physical and mental health, Wellbeing<br>COVID-19 Experiences and beliefs<br>Trait and State psychological measures<br>Open Ended Questions<br>Questionnaire evaluation Questions |
| 1st Follow-up Questionnaire (April 2021-June 2021) (n=5632) | Ethnicity<br>Nationality, religion and languages<br>Other demographics and education<br>Work<br>Home and Social Life<br>Harassment and discrimination<br>Physical and mental health, Wellbeing<br>COVID-19 Experiences and beliefs<br>Long COVID<br>Vaccine symptoms<br>Trait and State psychological measures<br>Open Ended Questions<br>Questionnaire evaluation Questions |
| 2nd Follow-up Questionnaire (October 2021-November 2021) (ongoing) | Ethnicity<br>Nationality, religion and languages<br>Other demographics and education<br>Work<br>Home and Social Life<br>Harassment and discrimination<br>Physical and mental health, Wellbeing<br>COVID-19 Experiences and beliefs<br>Long COVID |
|  | Vaccine symptoms<br>Trait and State psychological measures<br>Open Ended Questions<br>Questionnaire valuation Questions |
| Saliva Testing (Planned for October 2021) | DNA Data |
| Linkage to Electronic Healthcare Records (ongoing) | COVID-19 Clinical Outcomes (Acute infection, antibody status)<br>Comorbidities<br>Patterns of healthcare usage. |

In addition, participants gave permission to use data from their EHRs for a period of 25 years from the date of consent, allowing for longitudinal tracking of the effect of the pandemic on participants’ health. Not all participants who consented to have their EHRs linked completed the baseline questionnaire due to the consent process and the questionnaires being discrete options for participants.

Consented participants were asked to complete follow-up questionnaires 6 months (21^st^ April-26^th^ June 2021) and 10 months (18th October-26^th^ November 2021), after the study opened for participants. These repeated topics from the baseline questionnaire, with minor adjustments to reflect the changes in the COVID-19 pandemic in the UK. Due to the unique pressures that the COVID-19 pandemic has placed upon healthcare workers, limiting the amount of time available for participants to complete questionnaires, each questionnaire was designed so that it could be either standalone or be used in a longitudinal arrangement.

Between 18^th^ October – 26^th^ November 2021, participants were invited to provide consent to be sent a saliva sample kit, in order to collect DNA data. The samples will be stored at the UK Biocentre (Milton Keynes, UK) after initial processing.

Table 3 provides an overall summary of the data available from the cohort, while additional information on the questions asked, response options and question sources can be found in the UK-REACH data dictionary (https://www.uk-reach.org/data-dictionary).

## What has it found?

Table 4 shows age, ethnicity and sex of those in the cohort compared to the age distribution of those in the NHS in England (7). The cohort shows a very similar age distribution to the NHS workforce, with an average age differential of one year (8). While date of birth was captured during consent, ethnicity and sex were only captured by those who answered the baseline questionnaire, leading to variability in the amount of demographic information available from the cohort. The ethnicity information available however, demonstrates the UK-REACH cohort is more ethnically diverse than the NHS workforce with 26.6% of the UK-REACH cohort reporting a non-white ethnicity compared with 22.1% of the NHS workforce(9). Nevertheless, the study recruited fewer HCWs from white, Black and other ethnic groups than are present in the NHS workforce, whilst over-recruiting participants from Asian and mixed ethnic backgrounds. The UK-REACH cohort has a very similar sex balance as the NHS workforce, with 77% of the cohort female compared to 75.2% of the NHS (7). Data collected from the cohort have been used to produce multiple outputs, providing insight into HCWs during the COVID-19 pandemic, and a full list of these can be found at https://www.uk-reach.org/publications.

**Table 4:** Demographics Breakdown for the UK-REACH Cohort in comparison to the NHS Workforce.

| **Variable** | **UK-REACH**<br>**Cohort (1)**<br>**(n=17981)** | **NHS**<br>**Workforce (8)** |
| --- | --- | --- |
| **Age (years) (%)** |  |  |
| Under 25 | 3 | 6 |
| 25-34 | 23 | 23 |
| 35-44 | 25 | 24 |
| 45-54 | 27 | 28 |
| 55-64 | 18 | 18 |
| 65+ | 3 | 2 |
| **Sex (%) (n=15119)** |  |  |
| Male | 24.6 | 23 |
| Female | 75.2 | 77 |
| Prefer to use alternative term | 0.1 | N/A |
| Prefer not to answer | 0.1 | N/A |
| **Ethnicity (%) (n=15119)** |  |  |
| White | 61.1 | 77.9 |
| Black | 3.9 | 6.5 |
| Asian | 17.2 | 11.3 |
| Mixed | 3.7 | 1.9 |
| Other | 1.9 | 2.6 |
| Prefer not to answer/ Not available | 12.3 | N/A |

### Vaccine Hesitancy in HCWs

An analysis of the drivers of vaccine hesitancy was the first major finding from the UK-REACH cohort (10). This analysis of interim data collected from 4^th^ December 2020 to 19^th^ February 2021 included 11584 HCWs, of whom 23% reported vaccine hesitancy. HCWs from Black Caribbean (54.2% reported hesitancy), Mixed white and Black Caribbean (38.1%), Black African (34.4%), Chinese (33.1%), Pakistani (30.4%), and white Other (28.7%) ethnic groups were significantly more likely to be hesitant when compared to white British HCWs (21.3% hesitant). The following factors were also significant in predicting hesitancy towards the COVID-19 vaccine: younger age, female sex, higher score on a COVID-19 conspiracy beliefs scale, lower trust in employer, lack of influenza vaccine uptake in the previous season, previous COVID-19, and pregnancy. Qualitative analysis of a smaller selection of HCWs (n=99) from a separate work package of the UK-REACH project, who participated in face-to-face interviews and focus groups revealed a range of reasons that HCWs were hesitant about the COVID-19 vaccines. Reasons provided as contributors to vaccine hesitancy included: lack of trust in government and employers, safety concerns due to the speed of vaccine development, lack of ethnic diversity in vaccine studies, and confusing and conflicting information. Qualitative analysis also provided some strategies for addressing vaccine hesitancy in ethnic minority HCWs, such as inclusive communication, involving HCWs in the vaccine rollout, and promoting vaccination through trusted networks.

### Infection Risk in HCWs

HCWs, particularly those from ethnic minorities, have been shown to be at higher risk of infection with SARS-CoV-2 than the general population, although evidence is conflicted about the predictors and mediating factors of infection in HCWs (11). Analysis of 10 772 HCWs who reported working during the first UK national lockdown in March 2020 revealed that 2496 (23.2%) had some evidence of previous SARS-CoV-2 infection (via PCR tests, serology testing or self-reported COVID-19 diagnosis). Statistical analyses of the baseline UK-REACH survey revealed that demographic factors such as younger age and high religiosity were associated with an increased infection risk. A range of occupational factors were also associated with increased infection risk: attending to a higher number of SARS-CoV-2 positive patients, working in a nursing role (compared to a medical role), lack of access to PPE and working in an ambulance setting. HCWs working in an intensive care unit and those that worked in the south east of England or Scotland were at lower risk of infection (when compared to the West Midlands of England as a reference group). Black ethnic groups were initially identified as being at higher risk, but adjusted statistical models revealed factors which mediated the elevated infection risk (12).

### Personal Protective Equipment Access for HCWs

Access to personal protective equipment (PPE) may prevent transmission of SARS-CoV-2, and anecdotal reports exist of a lack of access to PPE by HCWs (13, 14). Two analyses were undertaken to examine the factors relating to PPE access for HCWs in the UK. The primary analysis included participants who answered baseline questions about access to PPE during the first UK national lockdown (23^rd^ March 2020) (n=10 508), while the secondary analysis included those who answered baseline questions about PPE access during the baseline questionnaire period (4^th^ December 2020-28^th^ February 2021) (n=12 252). The primary analysis found that only 35.2% of HCWs reported being able to access appropriate PPE all of the time during the first UK national lockdown, while the secondary analysis found that 83.9% of HCWs had access to PPE all of the time during the baseline questionnaire period (15). Several factors predicted access to PPE in both analyses, such as age (being older predicted greater access to PPE), being Asian (vs white), and role (allied health professionals, dentists and those who saw the most COVID-19 patients were all predictors of reduced access to PPE all the time). Both analyses also showed that access to PPE was not uniform across the UK, as those in South West and North West England were able to access PPE more frequently than those in London. In summary, access to PPE for HCWs was particularly limited during the first lockdown, and access varied based on sociodemographic, occupational and geographic factors (15).

## What are the main Strengths and Weaknesses?

The UK-REACH study is a UK-wide study, capturing information from the wide range of roles that form healthcare services in the UK, including ancillary workers who are often not included in such studies. The involvement of the healthcare regulators, NHS trusts and health boards, and various membership bodies has enabled the study to recruit from a large pool of HCWs, providing a diverse and representative sample of the wide range of clinical-based job roles within the UK healthcare sector. However, some staff such as porters, cleaners and kitchen staff are under-represented in the cohort, despite a targeted approach to recruit from these groups in collaboration with Serco, a UK public services provider, who are routinely contracted to provide ancillary staff in healthcare sites across the UK. The lack of representation in the cohort from groups with lower socioeconomic status may cause findings to under report the effects of outcomes on those groups.

A significant strength of the UK-REACH study is the ethnic diversity of the cohort, with 26.6% identifying with an ethnic minority background. Particularly as ethnic minorities are often underrepresented in studies (16). Nevertheless, Black ethnic groups remain under-represented in the UK-REACH study, which should be a key target for future studies of both COVID-19 and HCW occupational health with learnings from the UK-REACH study made available to facilitate this. In future, the high percentage of ethnic minority HCWs present in the cohort will allow for wider research questions to be asked, outside the current COVID-19 focus.

It is likely that the effects of the pandemic have placed additional strains on HCWs of all ethnicities for an extended period, which may have limited study participation, possibly because participants do not have time or do not wish to answer large numbers of questions about how the pandemic has affected them.

The online-only nature of the UK-REACH study enabled recruitment of participants across the UK, giving a national picture of the impact of the COVID-19 pandemic on HCWs of varying ethnicities. However, the exclusive use of digital communication methods (e.g. email and social media) to advertise the study, and digital data collection may have limited participation in the study, particularly amongst certain staff groups, such as those without access to a computer routinely throughout. As result the study is likely to contain biases due to participant self-selection. Initial recruitment was maximised via repeated communications from healthcare regulators and NHS trusts, with many participants receiving invitations to participate from both their regulator and their employer at different times. Reminder emails were also sent by the study team to participants who had registered their interest with the study by creating an account on the study website, but had not completed the consent process, and to participants who had consented to the study, but had not completed the baseline questionnaire. For the follow up questionnaires, consented participants were contacted to invite them to fill in the questionnaires, with reminders to participants to encourage completion.

## Can I get hold of the data? Where can I find out more?

The cohort website (https://www.uk-reach.org) contains an up-to-date record of all research activities, including publications in peer-reviewed journals, pre-print articles and other related study news.

Participants have consented to their pseudonymised data being made available to other approved researchers, and we welcome requests for collaboration and data access. Access to the resource requires completion of a proposal form, including a lay summary of the proposed research. Applications to access the resource will be assessed for consistency with the data access policy by the Scientific Committee, which has participant representation. Interested researchers are encouraged to contact the study management team via. Access to forms and more detail on the process can be found at https://www.uk-reach.org/data_sharing.

## Data Availability

Request data access and collaborations by following documentation found at https://www.uk-reach.org/main/data_sharing.

https://www.uk-reach.org/main/data-sharing

## Ethics Approval

The Study was approved by the Health Research Authority (Brighton and Sussex Research Ethics Committee; ethics reference: 20/HRA/4718). All participants gave electronic, informed consent. Trial ID: ISRCTN11811602.

## Acknowledgements

We would like to thank all the participants who take part in this study when the NHS is under immense pressure. We wish to acknowledge the Professional Expert Panel group (Amir Burney, Association of Pakistani Physicians of Northern Europe; Tiffanie Harrison; London North West University Healthcare NHS Trust; Ahmed Hashim, Sudanese Doctors Association; Sandra Kazembe, University Hospitals Leicester NHS Trust; Susie M. Lagrata (Co-chair), Filipino Nurses Association-UK & University College London Hospitals NHS Foundation Trust; Satheesh Mathew, British Association of Physicians of Indian Origin; Juliette Mutuyimana, Kingston Hospitals NHS Trust; Padmasayee Papineni (Co-chair), London North West University Healthcare NHS Trust),, the Steering and Advisory Group (1), Serco, as well as the following people for their support in setting up the study from the regulatory bodies: Kerrin Clapton and Andrew Ledgard (General Medical Council), Caroline Kenny (Nursing and Midwifery Council), David Teeman and Lisa Bainbridge (General Dental Council), My Phan and John Tse (General Pharmaceutical Council), Angharad Jones (General Optical Council), Katherine Timms and Charlotte Rogers (The Health and Care Professions Council) and Mark Neale (Pharmaceutical Society of Northern Ireland).

We would also like to acknowledge the following trusts and sites who recruited participants to the study: Affinity Care, Berkshire Healthcare NHS Trust, Birmingham and Solihull NHS Foundation Trust, Birmingham Community Healthcare NHS Foundation Trust, Black Country Community Healthcare NHS Foundation Trust, Bridgewater Community Healthcare NHS Trust, Central London Community Healthcare NHS Trust, Chesterfield Royal Hospital NHS Foundation Trust, County Durham and Darlington Foundation Trust, Derbyshire Healthcare NHS Foundation Trust, Lancashire Teaching Hospitals NHS Foundation Trust, Lewisham and Greenwich NHS Trust, London Ambulance NHS Trust, Northern Borders, Northumbria Healthcare NHS Foundation Trust, Nottinghamshire Healthcare NHS Foundation Trust, Royal Brompton and Harefield NHS trust, Royal Free NHS Foundation Trust, Sheffield Teaching Hospitals NHS Foundation Trust, South Central Ambulance Service NHS Trust, South Tees NHS Foundation Trust, St George’s University Hospital NHS Foundation Trust, Sussex Community NHS Foundation Trust, University Hospitals Coventry and Warwickshire NHS Trust, University Hospitals of Leicester NHS Trust, University Hospitals Southampton NHS Foundation Trust, Walsall Healthcare NHS Trust and Yeovil District Hospital NHS Foundation Trust.

## References

1. Woolf K, Melbourne C, Bryant L, Guyatt AL, McManus C, Gupta A, et al. The United Kingdom Research study into Ethnicity And COVID-19 outcomes in Healthcare workers (UK-REACH): Protocol for a prospective longitudinal cohort study of healthcare and ancillary workers in UK healthcare settings. medRxiv. 2021:2021.02.23.21251975.

2. Mutambudzi M, Niedzwiedz C, Macdonald EB, Leyland A, Mair F, Anderson J, et al. Occupation and risk of severe COVID-19: prospective cohort study of 120 075 UK Biobank participants. Occupational and Environmental Medicine. 2021;78(5):307–14.

3. Levene LS, Coles B, Davies MJ, Hanif W, Zaccardi F, Khunti K. COVID-19 cumulative mortality rates for frontline healthcare staff in England. Br J Gen Pract. 2020;70(696):327–8.

4. Nasir BW-SaR. Coronavirus (COVID-19) related deaths by occupation, England and Wales: deaths registered between 9 March and 28 December 2020: Office of National Statistics; 2021 [updated 25th Jan 2021.

5. Ayoubkhani D, Khunti K, Nafilyan V, Maddox T, Humberstone B, Diamond I, et al. Post-covid syndrome in individuals admitted to hospital with covid-19: retrospective cohort study. BMJ. 2021;372:n693.

6. Sahebi A, Nejati-Zarnaqi B, Moayedi S, Yousefi K, Torres M, Golitaleb M. The prevalence of anxiety and depression among healthcare workers during the COVID-19 pandemic: An umbrella review of meta-analyses. Prog Neuropsychopharmacol Biol Psychiatry. 2021;107:110247-.

7. Employers N. Gender in the NHS: NHS Employers; 2019 [Available from: https://www.nhsemployers.org/articles/gender-nhs-infographic.

8. Organisations N. Age in the NHS 2019 [cited 2021 27th August 2021]. Available from: https://www.nhsemployers.org/articles/age-nhs-infographic.

9. Digital N. NHS England Workforce Statistics - Ethnicity: NHS Digital; 2020 [updated 26th January 2021. Available from: https://www.ethnicity-facts-figures.service.gov.uk/workforce-and-business/workforce-diversity/nhs-workforce/latest.

10. Woolf K, McManus IC, Martin CA, Nellums LB, Guyatt AL, Melbourne C, et al. Ethnic differences in SARS-CoV-2 vaccine hesitancy in United Kingdom healthcare workers: Results from the UK-REACH prospective nationwide cohort study. The Lancet Regional Health - Europe. 2021:100180.

11. Evans S, Agnew E, Vynnycky E, Stimson J, Bhattacharya A, Rooney C, et al. The impact of testing and infection prevention and control strategies on within-hospital transmission dynamics of COVID-19 in English hospitals. Philosophical Transactions of the Royal Society B: Biological Sciences. 2021;376(1829):20200268.

12. Martin CA, Pan D, Melbourne C, Teece L, Aujayeb A, Baggaley RF, et al. Predictors of SARS-CoV-2 infection in a multi-ethnic cohort of United Kingdom healthcare workers: a prospective nationwide cohort study (UK-REACH). medRxiv. 2021:2021.12.16.21267934.

13. Liu M, Cheng S-Z, Xu K-W, Yang Y, Zhu Q-T, Zhang H, et al. Use of personal protective equipment against coronavirus disease 2019 by healthcare professionals in Wuhan, China: cross sectional study. BMJ. 2020;369:m2195.

14. Guardian T. Hancock:no evidence PPE shortages led to health and social care workers dying of Covid. The Guardian2021 [Available from: https://www.theguardian.com/politics/video/2021/jun/10/hancock-no-evidence-ppe-shortages-led-to-health-and-social-care-workers-dying-of-covid-video.

15. Martin CA, Pan D, Nazareth J, Aujayeb A, Bryant L, Carr S, et al. Access to personal protective equipment in healthcare workers during the COVID-19 pandemic in the United Kingdom: results from a nationwide cohort study (UK-REACH). medRxiv. 2021:2021.09.16.21263629.

16. Smart A, Harrison E. The under-representation of minority ethnic groups in UK medical research. Ethnicity & Health. 2017;22(1):65–82.

